# An ensemble multimodal approach for predicting first episode psychosis using structural MRI and cognitive assessments

**DOI:** 10.1101/2025.07.21.25331909

**Authors:** Sai Zhang

**Affiliations:** Institute of Psychiatry, Psychology & Neuroscience, King’s College London

**Keywords:** Schizophrenia, Ensemble Model, Multimodal, Data-driven

## Abstract

Classification between first episode psychosis (FEP) patients and healthy controls is of particular interest to the study of schizophrenia. However, predicting psychosis with cognitive assessments alone is prone to human errors and often lacks biological evidence to back up the findings. In this work, we combined a multimodal dataset of structural MRI and cognitive data to disentangle the detection of first-episode psychosis with a machine learning approach. For this purpose, we proposed a robust detection pipeline that explores the variables in high-order feature space. We applied the pipeline to Human Connectome Project for Early Psychosis (HCP-EP) dataset with 108 participants in EP and 47 controls. The pipeline demonstrated strong performance with 74.67% balanced accuracy on this task. Further feature analysis shows that the model is capable of identifying verified causative biological factors for the occurrence of psychosis based on volumetric MRI measurements, which suggests the potential of data-driven approaches for the search for neuroimaging biomarkers in future studies.

## 1 Introduction

Current psychiatric diagnostic systems such as DSM-5 and ICD-10 are mainly based on psychological and behavioral observations. Due to its dependency on the examination of psychiatric symptoms and signs, it is less associated with objective biological markers and other quantitive measurements. Agarwal et al. [1] argue that the absence of such criteria could lead to the ignorance of the causative factors. Therefore, recent studies focus more on the identification of genetic [2], biological [3] and biochemical [4] biomarkers of schizophrenia and try to establish a link in the diagnosis, treatment, and prognosis of the psychosis. Revealing such underlying factors can complement the lack of quantitive measurements and can potentially benefit the diagnosis and prognosis on the individual level. To rule out the influence of antipsychotics and the long-term effect of psychosis, Early Psychosis (EP), or First-Episode Psychosis (FEP) patients are of particular interest and have been widely used in the current literature.

Related studies have revealed the abnormalities including the structural and functional changes in cerebral regions for patients with schizophrenia through neuroimaging inspections. Alternations including cortical regions [5], sub-cortical regions [6] and neural connectivity changes [7] between the schizophrenia patients and healthy controls are stated to show observable differences. However, due to the complexity and heterogeneity of schizophrenia on an individual basis, challenges persist in fully utilizing the neuroimaging findings. Previous studies mainly focus on group-level feature differences [8] or work with the isolated brain regions [9], and the clinical translation is hard to even with significant outcomes in the experiments [10]. To tackle this problem, there is increasing interest in using machine learning algorithms as automatic feature extractors to improve the performance of structural imaging features. Xiao et al. [11] studied the classification of 163 first-episode patients and 163 healthy controls on cortical thickness and surface area features with a Support Vector Machine (SVM). Following similar methodologies, Guo et al. [12] selected features from amygdaloid and hippocampal regions and used SVM with additional sequential backward feature elimination. Later on, Yassin et al. [13] focused on the cortical thickness and subcortical volumes with a more complicated random forest classifier. These approaches primarily show the potential application of machine learning algorithms to identify neuroimaging biomarkers in a data-driven manner [14].

Although there have been efforts to classify schizophrenia patients with the controls, uncertainty still exists within these features and remains to be solved. Bearing these limitations in mind, Squarcina et al. [15] proposed using a model ensemble to provide a better estimation of the classification result. With SVM and multi-kernel learning techniques, it can tackle epistemic uncertainty through the design of different kernels. Moreover, Chilla et al. [16] further employed a multi-model approach together with a model ensemble on diverse neuroanatomical markers and demonstrated the strong potential of such an approach. The combination of multi-model data can provide complements of features from multiple sources and the ensemble of models can provide epistemic measurements of uncertainty quantification with different underlying models.

Cortical volumes and subcortical volumes extracted from Magnetic Resonance Imaging (MRI) are widely used in schizophrenia studies. The gyrification of the brain regions has been found to be related to the relative volume of these regions as well as the cognitive performance [17], which makes it suitable for a multi-model study together with cognitive assessments. In this study, we acquired data for patients and controls from the Human Connectome Project for Early Psychosis (HCP-EP) [18] including 108 schizophrenia patients (with affective and non-affective disorders) and 47 healthy controls. We examined brain region volumes together with the cognitive assessments to classify these two groups. The study shows that 1) model ensemble can provide better interpretation of the features with improved sensitivity and specificity, 2) multi-model data integration of cognitive and brain metrics serves as a complementary component to increase the overall prediction accuracy, and 3) machine learning methods can provide more insights on the biological evidence of first episode psychosis with the multimodal dataset. Our proposed processing pipeline is robust on a relatively small dataset with competitive performance against other works on a similar dataset. Additionally, the model ensemble approach can provide uncertainty quantification on the framework level, which is useful for providing case-by-case evaluation.

## 2 Methods

### 2.1 Subjects

The study included 108 participants with psychosis and 47 healthy controls. The availability of the dataset is publicly accessible via the National Institute of Mental Health Data Archive in accordance with the data usage policy. The diagnosis of psychosis is collected with Structured Clinical Interview for DSM-IV-TR [19].

The recruitment process of potential participants with psychosis relies largely on advertisements and in-person announcements within the community. For healthy control subjects, the advertisements are posted in the organizational newsletters and public facilities. From Table 1, it is clear that although the dataset is unbalanced between patients with first-episode psychosis and healthy controls, the distributions along sex, interview age, racial identity, and collection site are even.

**TABLE 1.**
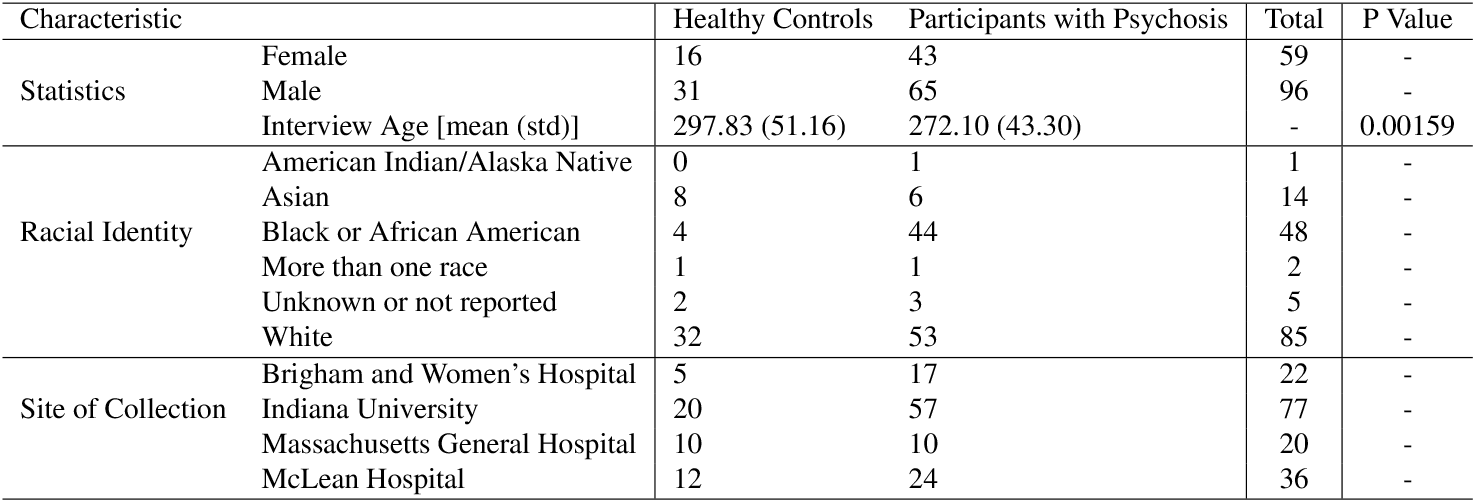
Clinical characteristics of first episode psychosis patients and healthy controls.

| Characteristic |  | Healthy Controls | Participants with Psychosis | Total | P Value |
| --- | --- | --- | --- | --- | --- |
| Statistics | Female | 16 | 43 | 59 | - |
|  | Male | 31 | 65 | 96 | - |
|  | Interview Age [mean (std)] | 297.83 (51.16) | 272.10 (43.30) | - | 0.00159 |
| Racial Identity | American Indian/Alaska Native | 0 | 1 | 1 | - |
|  | Asian | 8 | 6 | 14 | - |
|  | Black or African American | 4 | 44 | 48 | - |
|  | More than one race | 1 | 1 | 2 | - |
|  | Unknown or not reported | 2 | 3 | 5 | - |
|  | White | 32 | 53 | 85 | - |
| Site of Collection | Brigham and Women's Hospital | 5 | 17 | 22 | - |
|  | Indiana University | 20 | 57 | 77 | - |
|  | Massachusetts General Hospital | 10 | 10 | 20 | - |
|  | McLean Hospital | 12 | 24 | 36 | - |

### 2.2 Data Acquisition

For MRI data acquisition, structural scans are collected from the participants following the same setup as in the HCP Lifespan protocol [20]. The images are performed under 3 Tesla, using multi-echo T_1_*w* with variable flip angle and turbo-spin-echo T_2_*w* scans, as stated in HCP-EP project [18]. It is further processed by the FreeSurfer (v7.3.1) image analysis package (http://surfer.nmr.mgh.harvard.edu/). With the default parcellation and the Desikan atlas, we obtained 68 brain regions for volumetric measurements, which are used as the brain structural features for the machine learning pipeline. Before the next steps, the volume is normalized with the estimated intracranial volume to represent the volume in percentages.

For the cognitive assessments, we adopted several dimensions from the HCP-EP project [18]. Speed of processing is measured in multiple tests including the sweeps assessment from PositScience Corporation^1^. Emotion recognition dimensions are measured with the 40-item Penn Emotion Recognition Test (ER-40) [21] and the PROID test by PositScience Corporation. Six emotion recognition abilities including happiness, sadness, anger, fear, surprise, and disgust are assessed in these tests. Cognitive battery with NIH Toolbox [22] is performed. Additional features included tests for working memory, reading, fluency, and attention inhibiting under the stimulus.

### 2.3 Data Analysis

Given the inherent nature of multimodal datasets, we performed data analysis on the brain volumetric features and cognitive features separately. In the process of analysis, we aim to reveal the underlying distribution of the data which explains our choices of design for the classification pipeline.

For volumetric cortical and subcortical brain region measurements, the volume data is transformed into relative percentages with respect to the estimated intracranial volume. The correlation heatmap between the relative volumes of these regions is shown in Figure 1. From the graph, we can see that volume data collected from the right hemisphere (RH) has a strong correlation with the left hemisphere (LH) for both cortical and subcortical regions. This indicates the possibility of dimensionality reduction with no prior knowledge of classification types, which is adopted in our detection pipeline.

For cognitive assessments, we initially identified a small number of ER-40 emotional recognition features with invalid zero measurements for several cases. Therefore, we performed mean value replacement for these features. Then, we visualized the distribution of emotion recognition tests from ER-40 tests as illustrated in Figure 4.

**FIGURE 1:**
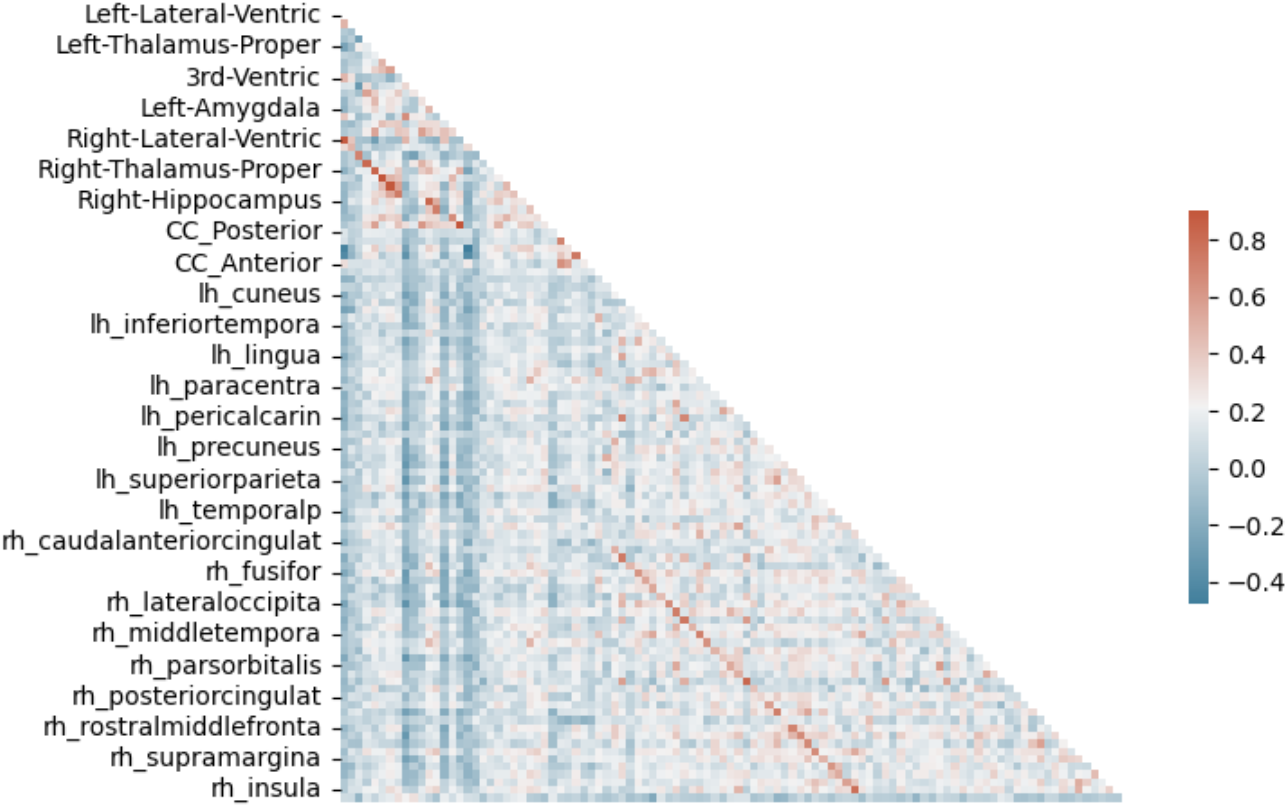
Brain region volume correlation from structural MRI

**FIGURE 2:**
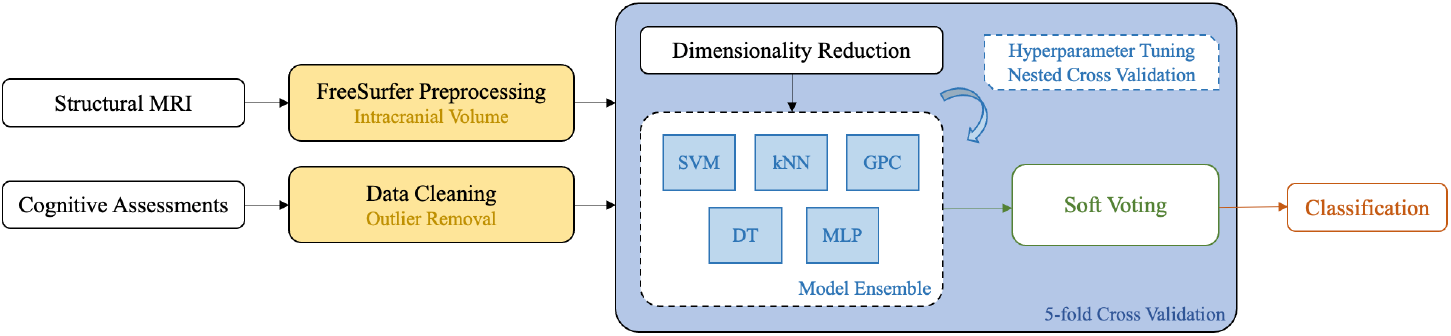
Proposed Classification Pipeline

**FIGURE 3:**
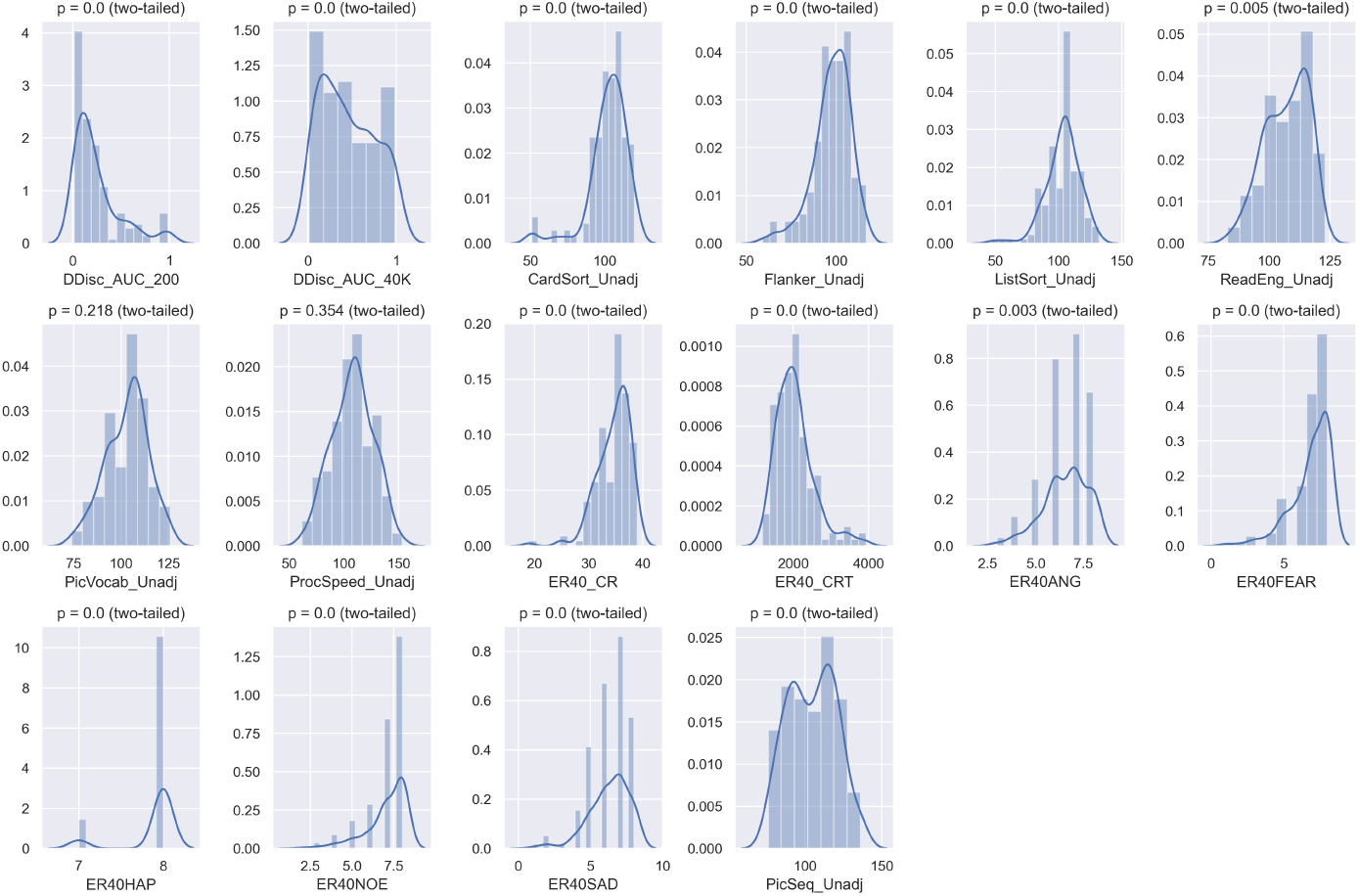
Cognitive assessment features distribution

**FIGURE 4:**
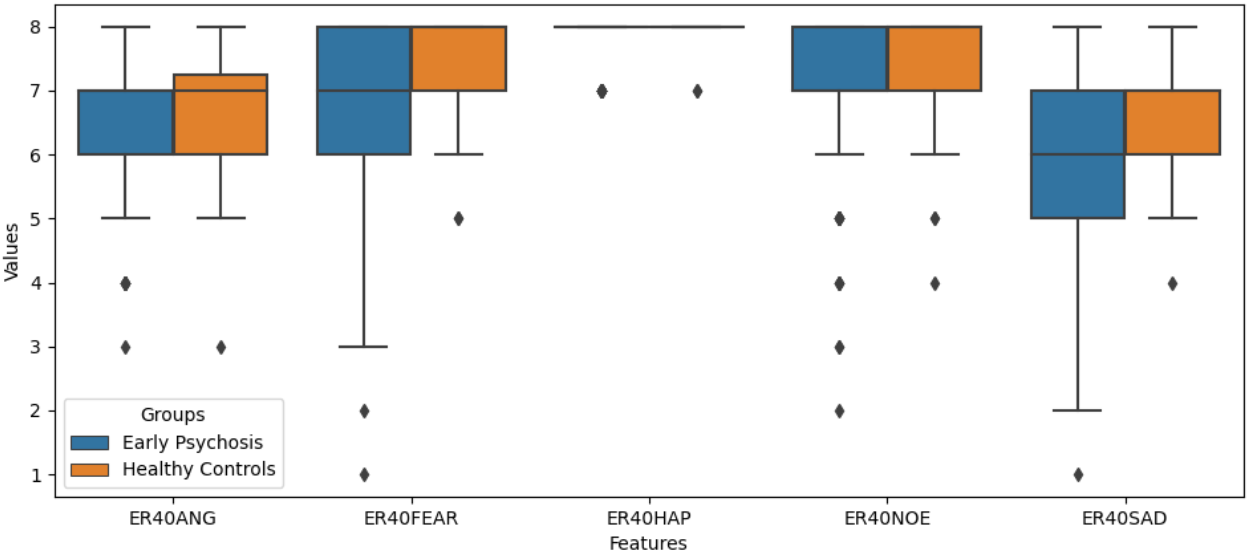
ER-40 emotion recognition features distribution

For both features collected, we assumed that the dataset follows gaussian distribution and tested the accordance with D’Agostino and Pearson’s normal test for skew and kurtosis, with the null hypothesis that a sample comes from a normal distribution. For the histograms and p-values of these features, please refer to Figure 7 in Appendix A.

In addition to statistical analysis of the features, we also performed a sensitivity analysis for feature importance given the phenotypes. The dependent analysis on classification labels is useful for the potential identification of influential factors [23]. We adopted a randomized decision tree to perform linear sensitivity analysis on the features from structural MRI and cognitive assessments. The bar chart for top important features is shown in Figure 5 and 6.

**FIGURE 5:**
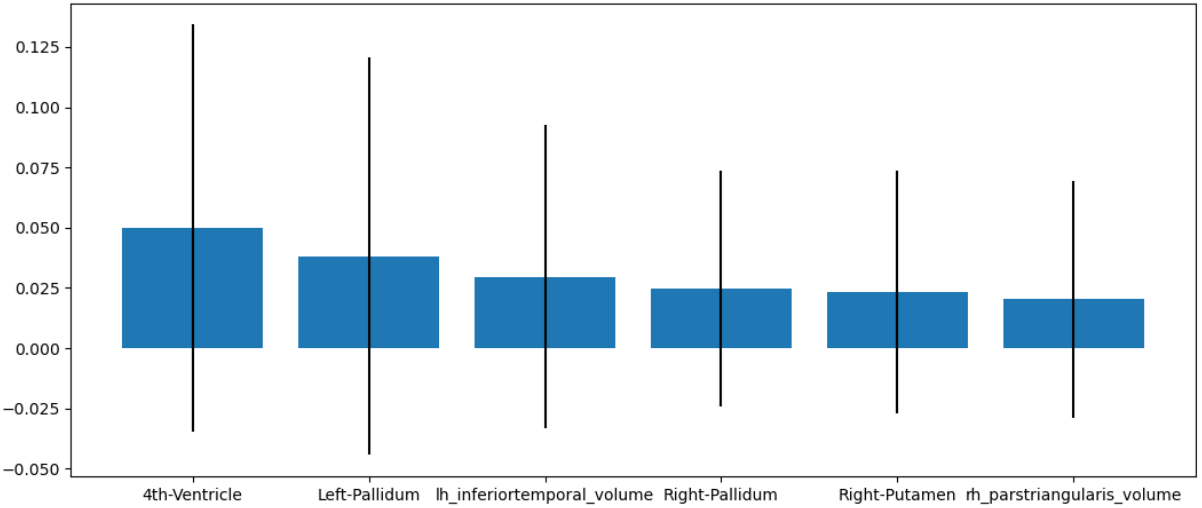
Top 6 important features with structural MRI from random decision tree

**FIGURE 6:**
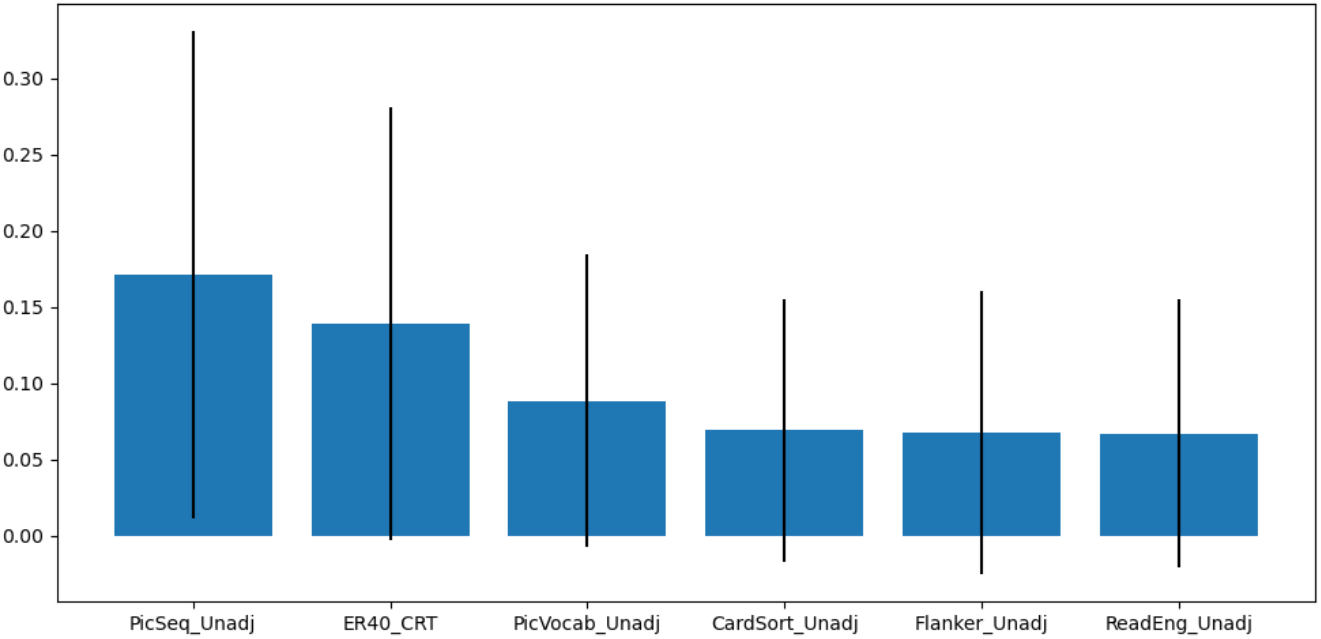
Top 6 important features with cognitive data from random decision tree

**FIGURE 7:**
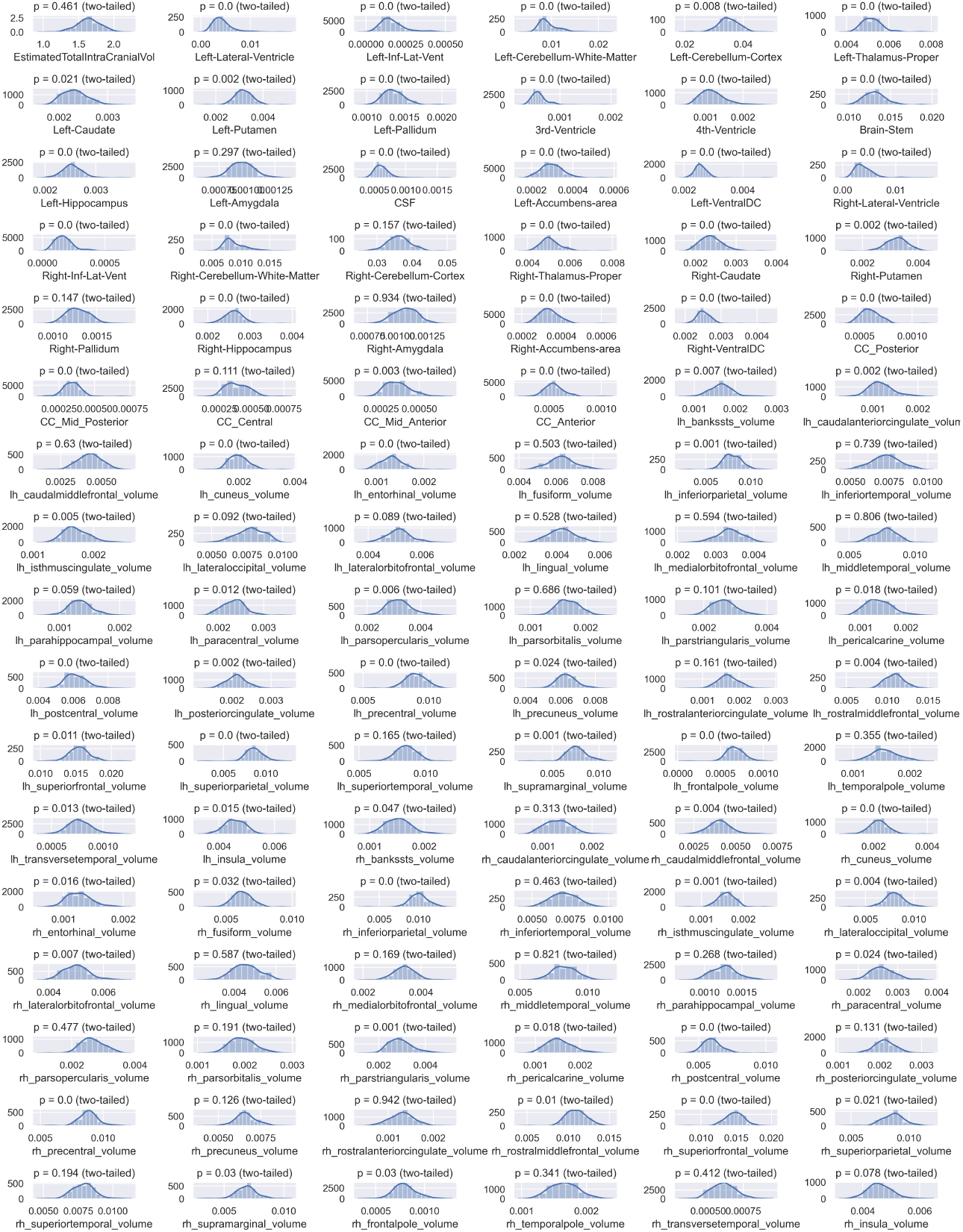
Structural MRI features distribution

Comparing these two multimodal datasets, we can see that the cognitive dataset has a better overall relationship with the psychosis predictions, with the top-1 feature Pic-Seq_Unadj determining 16.9% of the result while the top-1 volumetric measurement of 4th-Ventricle only explaining 5% of the differences. Although the feature importance statistics are derived from the decision tree which is a linear model, the analysis can still serve as a compared sensitivity analysis between the ensemble model approach and the traditional SVM approach, combined with further ablation tests in Section 3.2.

### 2.4 Classification Pipeline

Based on the initial data analysis, we designed a classification pipeline for the detection of first-episode psychosis. We adopted a nested-fold cross-validation scheme for model selection, hyperparameters tuning, and evaluation of our ensemble model. For the multimodal dataset, we first studied the performance with independent measures and then combined them to compare the efficacy of the multimodal approach. Major detailed steps of the pipeline are as below.

#### Data Preprocessing

For pre-processing stage, we carefully designed this process to avoid any correlation between features when using a cross-validation setup. Following the initial data processing in Section 2.3, we performed outlier detection on the entire dataset with IsolationForest. Then, all the transformations on the dataset are performed within each train and test split. The data is first to split into train and validation subsets with stratified 5-fold splitting. Since the distribution of the data is uneven, the stratified setup can benefit the stability of the training process. Within each split, we normalized the dataset to zero mean and unit variance with StandardScaler from the sci-kit-learn library. With the standardized dataset, we further reduced the dimensionality of the features with Principle Component Analysis (PCA) [24]. The PCA is independent of the predicted labels while preserving orthogonal features in lower dimensional latent space. For most of the classifiers, the curse of dimensionality is inevitable with increased uncertainty on samples and degraded performance. The data setup for model selection is slightly different, with nested cross-validation to validate the tuned parameters on the outer loop.

Model Selection: The model selection stage includes model fine-tuning and voting optimization. For model fine-tuning, we used 5-fold nested cross-validation, with inner training and testing set for hyper-parameter testing and outer validation set for performance assessments. To find the best set of parameter setups, we adopted a grid search strategy on parameter subspace with GridSearchCV. For the model zoo, we mainly considered the simplicity of the model as well as the different underlying mechanisms. The selected models include Multi-Layer Perceptron (MLP), k-Nearest Neighbor (kNN), Gaussian Process (GP), Support Vector Machine (SVM), and Decision Tree (DT) Classifier. The difference in decision boundary for these models can greatly benefit the model’s robustness and performance. For the voting stage, we used a soft voting strategy, with a similar nested cross-validation setup for model tuning. The final ensemble model is evaluated with averaged performance on the cross-validation dataset. An overview of the processing pipeline is shown in Figure 2.

#### Study Design

Given the multimodal dataset, the study is divided into two parts, including classification with independent measurements and classification with combined measurements. With independent measurements, we employ the pipeline for structural MRI and cognitive assessments separately. For combined results, we rerun the entire pipeline on inner-joined tables of multimodal features with redundant columns removed. Through the splitting and merging of the dataset, we aim to quantitively compare and argue the effectiveness of our pipeline design and show the advantage of our proposed structure.

#### Architectural Design

For easy implementation and reuse of all the components, we largely followed the infrastructures of the sci-kit-learn package. We designed the wrapper class for PCA embedded processing pipeline and combined it with a number of estimators from scipy. The cross-validation and nested cross-validation model is abstracted into a data loader, data estimator, and data analyzer, allowing a flexible combination of all the components. The open-source implementation of this work can be found on Github^2^.

## 3 Results

The quantitive performance analysis is covered in this chapter. In the performance section, we studied the overall classification result from the multimodal ensemble model and demonstrated the efficacy of employing such an approach. To study the importance of each feature in the ensemble model, we further adapted the ablation test on all of the features to determine the decision variables for the model. By comparing the feature importance with ranking in Section 2.3, we identified the major difference in the dominating features compared to a linear model, which explained the performance boost.

### 3.1 Performance

To quantitively study the classification outcome for first episode psychosis, we modeled the performance from multiple evaluation metrics. For accuracy consideration, we adopted precision and class-balanced accuracy. Due to the imbalanced nature of the dataset, we further calculated the specificity, recall, and f1-score of the classes.

#### Independent Measurements

For the analysis of the independent measurements, we fine-tuned model parameters based on the grid search as indicated in Table 5 and summarized the model performance on the volumetric data and cognitive data separately as shown in Table 2.

**TABLE 2.** Model Performance on Independent Measurements

| Measurement | Model | Weighted F1-Score |  | Balanced Accuracy |  | Accuracy |  | Sensitivity |  | Specificity |  |
| --- | --- | --- | --- | --- | --- | --- | --- | --- | --- | --- | --- |
|  |  | Mean | Std | Mean | Std | Mean | Std | Mean | Std | Mean | Std |
| Structural MRI | Support Vector Machine | 0.6709 | **0.0453** | 0.5952 | **0.0394** | **0.7285** | 0.0649 | 0.7190 | **0.0404** | 0.2485 | **0.0746** |
|  | Gaussian Process | 0.6495 | 0.0623 | 0.5725 | 0.0692 | 0.6886 | 0.0982 | 0.6906 | 0.0664 | 0.2439 | 0.0942 |
|  | k-Nearest Neighbor | 0.6157 | 0.0615 | 0.5812 | 0.0857 | 0.6347 | 0.0760 | 0.6122 | 0.0580 | **0.4924** | 0.1888 |
|  | Decision Tree | 0.6338 | 0.0654 | 0.5935 | 0.0530 | 0.6518 | **0.0515** | 0.6343 | 0.0760 | 0.4773 | 0.1048 |
|  | Multi-Layer Perceptron | 0.6541 | 0.0588 | 0.5898 | 0.0613 | 0.6702 | 0.0651 | 0.6733 | 0.0699 | 0.3530 | 0.1045 |
|  | Ensemble Model | **0.6919** | 0.0609 | **0.6173** | 0.0650 | 0.7273 | 0.0904 | **0.7246** | 0.0585 | 0.3167 | 0.0939 |
| Cognitive Assessment | Support Vector Machine | 0.7178 | **0.0301** | 0.6573 | **0.0502** | 0.7492 | 0.0560 | 0.7343 | 0.0370 | 0.4600 | 0.1824 |
|  | Gaussian Process | 0.7170 | 0.0303 | 0.6498 | 0.0505 | 0.7495 | **0.0443** | 0.7407 | **0.0208** | 0.4178 | 0.1795 |
|  | k-Nearest Neighbor | 0.6929 | 0.0854 | 0.6713 | 0.0772 | 0.7433 | 0.0886 | 0.6901 | 0.0969 | **0.6244** | 0.1805 |
|  | Decision Tree | 0.6800 | 0.0676 | 0.6209 | 0.0817 | 0.6822 | 0.0718 | 0.6839 | 0.0669 | 0.4600 | 0.1467 |
|  | Multi-Layer Perceptron | 0.7341 | 0.0696 | 0.6749 | 0.0615 | 0.7480 | 0.0791 | 0.7409 | 0.0752 | 0.5044 | **0.1008** |
|  | Ensemble Model | **0.7432** | 0.0471 | **0.6818** | 0.0525 | **0.7612** | 0.0647 | **0.7534** | 0.0528 | 0.5000 | 0.1229 |

From the table, we can see that the most frequently used Support Vector Machine (SVM) performs moderately well on these two measurements, with relatively small standard variation and top accuracy. Overall, the predictions made by volumetric structural MRI data are weaker than those using cognitive counterparts. This result can be inferred from the feature importance analysis, where the indicator for the former is significantly smaller than the latter. If we ignore the ensemble model, the top-performing model for structural MRI is the SVM with 67.09% F1-Score while for the cognitive dataset is Multi-Layer Perceptron (MLP) with 73.41% F1-Score. In these two datasets, the ensemble model demonstrated a strong performance boost compared to using any one of these models alone. The ensemble model on structural MRI data achieved 69.19% on F1-Score with > 50% soft voting policy. Likewise, the model shows a 74.32% F1-Score for cognitive assessments with the same voting strategy. The performance gap between the ensemble model and separate ones is significant by more than 1%, which indicates that the underlying mechanism for different models does account for the epistemic uncertainty for the detection of first-episode psychosis.

#### Combined Measurements

For combined measurements, we performed a similar analysis as for the previous section, and the fine-tuned parameters are shown in Table 5. The quantitive results are listed in Table 3. Compared with independent measurements, most of the models suffer from the additional dimensionality of the input dataset even with the PCA dimension reduction technique. However, the ensemble model outperforms all the previous attempts by a large 4% margin compared to the combined SVM approach and achieved 77.07% on F1-Score. The decision strategy for the ensemble model uses > 60% soft voting to eliminate uncertain cases. From the results, we can see that the combined dataset does provide additional information to the model and the cognitive assessments and volumetric brain data serves as complementary to each other.

**TABLE 3.** Model Performance on Combined Measurements

| Model | Weighted F1-Score |  | Balanced Accuracy |  | Accuracy |  | Sensitivity |  | Specificity |  |
| --- | --- | --- | --- | --- | --- | --- | --- | --- | --- | --- |
|  | Mean | Std | Mean | Std | Mean | Std | Mean | Std | Mean | Std |
| Support Vector Machine | 0.7322 | 0.0512 | 0.6662 | **0.0356** | 0.7619 | 0.0589 | 0.7484 | 0.0626 | 0.4644 | 0.1431 |
| Gaussian Process | 0.6657 | **0.0075** | 0.6311 | 0.0523 | 0.6991 | **0.0444** | 0.6710 | **0.0241** | 0.5311 | 0.2352 |
| k-Nearest Neighbor | 0.6962 | 0.1109 | 0.6843 | 0.0895 | 0.7366 | 0.0782 | 0.6903 | 0.1165 | 0.6778 | 0.1052 |
| Decision Tree | 0.7020 | 0.0676 | 0.6271 | 0.0564 | 0.7100 | 0.0774 | 0.7161 | 0.0747 | 0.4044 | **0.0752** |
| Multi-Layer Perceptron | 0.6982 | 0.0969 | 0.6518 | 0.0957 | 0.7265 | 0.0760 | 0.7161 | 0.0966 | 0.4911 | 0.2396 |
| Ensemble Model | **0.7707** | 0.1316 | **0.7467** | 0.1412 | **0.7966** | 0.1124 | **0.7742** | 0.1338 | **0.6822** | 0.2362 |

Although we only used hard 0/1 classification classes for healthy controls and participants with first-episode psychosis when training and evaluating the model, we can still measure the epistemic uncertainty based on the trained model. The soft voting strategy in the ensemble model collects predictions from a bag of models and the collected value can be treated as the confidence of the overall prediction outcome. With an averaged predicted output equal to one, it indicates that all the models agreed firmly on the positive sign of psychosis while a value closer to 1/2 might indicate high uncertainty in the belonging of the sample for a variety of methods. Uncertainty measurements, especially in a case-by-case manner for the patients, are of great importance to the clinical application of the algorithm.

### 3.2 Feature Importance Analysis

To study the dominating factors for the combined model, we employed an ablation test on each feature in the combined dataset. To identify such features, we can determine which input column affects the prediction outcome of the model by a relatively large amount if the given feature is completely removed from the scope. In this test, we used the same parameter setup for the multimodal ensemble model, initialized the model in a 5-fold cross-validation scheme, and followed the same data preprocessing steps. For stability and reproducibility concerns, we duplicated the ablation for each feature by 10 times and collected the average reported accuracy for reversed ranking of important features.

The top 10 ranked features in both cohorts are listed in Table 4. Compared to the feature importance analysis made by linear models, the results are slightly different than the ensemble approach. For cognitive assessments, the top-ranked correct response time and picture sequence memory test is surpassed by the emotion recognition test of anger and fear. This result indicates that the incorrect response to the detection of fear and anger may act as a strong indicator of psychosis. For volumetric measurements, the volume of the temporal pole in the left hemisphere and right hemisphere both show up as a strong influencer in the prediction, followed up by the supramarginal gyrus and transverse temporal gyrus. Although the importance of these factors differs greatly in the linear decision tree model, the performance degradation of removing these features in our ensemble model is very close to each other.

**TABLE 4.** Averaged Performance Degradation on Feature Ablation Test

| Ranking | Feature | Averaged Balanced Accuracy Degradation (%) |
| --- | --- | --- |
| 1 | ER40ANG | 3.7009 |
| 2 | ER40FEAR | 3.7007 |
| 3 | ER40_CR | 3.6992 |
| 4 | ProcSpeed_Unadj | 3.6972 |
| 5 | lh_temporalpole_volume | 3.6960 |
| 6 | rh_temporalpole_volume | 3.6956 |
| 7 | ReadEng_Unadj | 3.6951 |
| 8 | rh_supramarginal_volume | 3.6943 |
| 9 | ER40_CRT | 3.6939 |
| 10 | rh_transversetemporal_volume | 3.6929 |

**TABLE 5.**
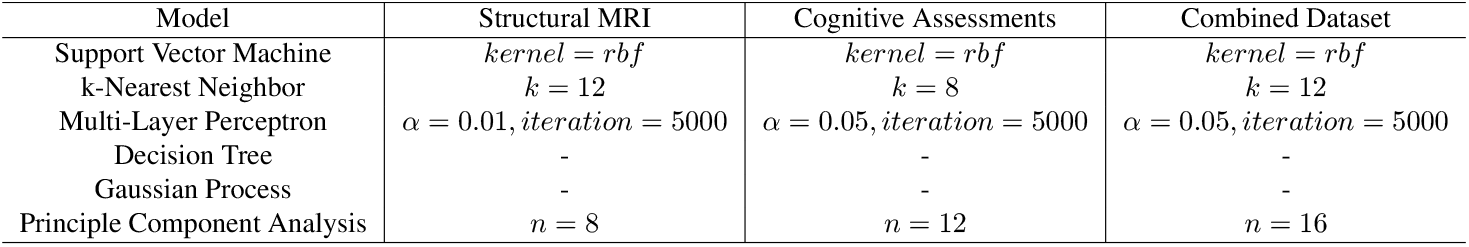
Fine-tuned Model Parameters

| Model | Structural MRI | Cognitive Assessments | Combined Dataset |
| --- | --- | --- | --- |
| Support Vector Machine | $kernel = rbf$ | $kernel = rbf$ | $kernel = rbf$ |
| k-Nearest Neighbor | $k = 12$ | $k = 8$ | $k = 12$ |
| Multi-Layer Perceptron | $\alpha = 0.01, iteration = 5000$ | $\alpha = 0.05, iteration = 5000$ | $\alpha = 0.05, iteration = 5000$ |
| Decision Tree | - | - | - |
| Gaussian Process | - | - | - |
| Principle Component Analysis | $n = 8$ | $n = 12$ | $n = 16$ |

Part of the reason for this difference in feature utilization could be introduced by different underlying decision boundaries for the models in the ensemble pipeline. The combination of models enables the framework to identify such features and interprets them properly. Since our approach performs significantly better than the naive SVM model, it is worth investigating this gap with the differences in feature interpretation.

Interestingly, although we can instantly identify the happiness measurement from the cognitive assessments as an abnormal distribution in Figure 4, its feature importance in the model trained with an uncleaned dataset is ranked #10 among all the other measurements. This indicates that the happiness measurement, though as odd as it seems, might still be reliable under a small volume of the dataset.

## 4 Discussion

In this study, we aimed to discuss the feasibility of endorsing a multimodal dataset with an ensemble model to distinguish first-episode psychosis participants apart from healthy controls. By adopting a robust yet elegant pipeline, we can move the same pipeline for datasets on different cohorts and demonstrate competitive performance with similar works. Furthermore, the ensemble model approach is proven to be useful in a multimodal scenario, given that the performance boost is significant compared to using a single model alone. Apart from performance considerations, our proposed approach also features epistemic uncertainty quantification, which is a rising need for clinical applications for machine learning models as a complementary interpretability layer of the predictions.

In addition to do predictions on the given dataset, we also tried to identify important features from the dataset through the integration of modalities. Known as an automatic feature extractor [25], these classifiers are capable of generating decision boundaries to distinguish the presence of psychosis. Through the analysis of the decision made by these models, it is possible to generate new insights into these features in a data-driven way. In our work, we demonstrated the feature importance rankings both from linear models and from our processing pipeline. The comparison between them can not only explain the main difference in the performance boost of our pipeline but also allow us to interpret the dataset. Although we used cognitive data and brain volumetric data from different cohorts, we can establish strong links between cognitive assessments and biological evidence. For the volumetric information on the temporal pole, earlier findings [26] indicated its preferential function in complex visual scene analysis including face recognition and visual memory, which is later appraised by Herlin et al. [27] recently. In our findings, the temporal pole in both the left and right hemispheres are all among the top-ranked features, which correspond to the high-ranked emotional tests in cognitive assessments. The volume of the supramarginal gyrus in the right hemisphere also corresponds to the cognitive features. In a recent study, Wada et al. [28] summarized the conclusion with path analysis and emotion recognition scores that the volume of the right supramarginal gyrus is associated with emotion recognition ability, which agrees with the analysis by our model. As for the transverse temporal region, it is found to be related to auditory functionalities [29]. Related studies by Ohi et al. [30] spotted structural alterations in the same area for patients with schizophrenia, and authors in [31] found that emotional intensified pictures can enhance the auditory novelty processing in this region. All of the biological features identified by our method are confirmed to be related to the brain functionality of emotion recognition, visual memory, and auditory ability, while most of them are related to emotional abilities. This finding shows the importance of emotion-related tests in psychosis assessments, with other biological pathways, remaining to be explored.

The strength of our work can be summarized below. To begin with, the relatively high-performance scores across multiple metrics indicate the usefulness of the binary predictions. Second, the uncertainty quantification functions that come with the ensemble model approach are also preferable for clinical applications with case-by-case statistics. Further analysis shows that features identified by our model are known to be informative for brain cognitive functionalities which might be the causative factor for the presence of first-episode psychosis. Our result shows that machine learning techniques can help provide an individualized diagnosis with high confidence, disentangling complex features, adding interpretability to the model, and serving as biological indicators for future studies. Despite the promising outcome of this work, several limitations need to be addressed.

First, we conducted our experiments with a moderately sized dataset, where the generality of the outcome remains questioned. The generality of the model on cross-site measurements is critical for clinical transitions and is worth investigating. Second, the gap between the systematic diagnosis of the psychosis and the underlying causes remains hidden due to the complex causation factors. Although we unveiled part of the connections between cognitive and structural MRI features, the majority of the connections remain unexplored. Third, we excluded the subtypes of first episode psychosis from the scope of the model, which includes more hints on the psychopathology of the illness. Possible directions for future studies include the combination of a larger dataset in more modalities, thorough reasoning behind the selection of ensemble kernels, and more exploration of the feature analysis part. As suggested in this work, the promising relationships found in the identified critical features can be further studied following the data-driven approach, with the possibility to reveal new links on individualized biological factors of first episode psychosis.

## Data Availability

All data produced in the present study are available upon reasonable request to the authors.

https://github.com/Catherine9811/HCP-EP-Codebase

## Acknowledgements

I was originally allocated to Prof. Nikolaos to extend the functionality of NeuroMiner. I want to express my gratitude to him as well as Dr. Dong for his help in the initial months. Due to some reasons, I am unable to continue my project with Nikolaos. Here, I must thank Dr. Sandra for her help as my supervisor. She is always supportive, and responsive and encouraged me to explore in a wide context. I couldn’t imagine how I can finish my project on time without you.

Many thanks to all the lecturers in Early Intervention of Psychosis. The broad content of the course greatly inspired me to extend my research, and the materials are of great importance to my future career.

## Appendices

### A Dataset Distribution

### B Model Parameters

## Footnotes

1 Brain HQ: https://www.brainhq.com

2 HCP-EP Codebase: https://github.com/Catherine9811/HCP-EP-Codebase

